# Assessment of Adverse Events Reported to The Poison Control and Forensic Chemistry Center in Jeddah, Saudi Arabia

**DOI:** 10.1101/2020.04.27.20077420

**Authors:** Nawaf Almuntashiri, Osama Alsahafi, Mohammad Gamaruddin, Sherif Attia Hammad, Mansour Tobaiqy

## Abstract

**Background:** Poisoning is a major public health threat in many countries across the globe, including Saudi Arabia. Poison control facilities help to provide immediate treatment to the patients affected by poisoning. Poisoning cases from various regions of the Kingdom are reported to the Poison Control and Forensic Chemistry Centers.

**Aims:** This study aimed at assessment of the demographics, risk factors and management of various poisoning cases reported at the Poison Control and Forensic Chemistry Center in Jeddah, Saudi Arabia.

**Methods:** Data was collected from the poisoning archived forms, between 01-01-2019 to 30-12-2019. A data collection sheet was designed to collect information about the region of call, age and gender of the patient, type, place, route and pattern of poisoning. All the data was analyzed through SPSS software.

**Results:** Most cases of intoxications were occurred and reported from the Western Region of Saudi Arabia (n=97, 38%), and were for males (n=137, 54%). The majority of poisoning cases occurred in children under 5-years of age (n=198, 78%). Poisoning cases were accidental (n=233, 92%) and intentional (n=11, 4%), and most (n=246, 96%) occurred at home. Drug poisoning was more common (n=155, 61%) than chemical poisoning (n=92, 36%). The rout of poisoning was oral in the vast majority of incidents (n=242, 94%). Intentional poisoning was more common in females (n=8, 3%). All poisoning reports initiated by physicians and other healthcare professionals (n=253, 100%), within one hour of the poisoning (n=144, 57%) and after one home (n=109, 43%). Decontamination with active charcoal (n=62, 24%) was the most common method recommended to manage intoxicated patients, followed by gastric lavage (n=9, 3.5%).

**Conclusion:** The current study assessed the reports of adverse events in a poisoning center in Saudi Arabia, most cases were related to medicines, followed by chemicals and most occurred at home. Most of poisoning cases are unintentional and occurred at home due to ingestion of medicinal products. Therefore, awareness of parents about the potent poisons at home may help to minimize the occurrence of such adverse events.

## Introduction

Poisoning represents an injury caused due to exposure to an extrinsic compound which leads to cell injury and/or death. Exposure to poisons can be via absorption, ingestion, inhalation, or injection. The exposure can be acute or chronic, causing variable clinical presentations. Factors establishing the severity and outcomes of poisoning in an individual are interlinked and comprise dose, type of poison, formulation, route, presence of other toxins, and presence of other injuries^(1–2)^. children are at greater risk of accidental poisoning ^(2)^. The cases of accidental poisoning are quite common in the young ones due to innate enthusiasm to discover their surroundings with their taste^(2)^.

Poisoning incidents are a major health problem around the globe, the magnitude of which varies from one country to another. The data from the WHO reported that in 2002, roughly 350,000 people died due to accidental poisoning while in 2004, more than 45,000 deaths in individuals aged above 20 years died of acute cases of poisoning. It is also the 2^nd^ common reason for injury and hospitalization in children aged below 5 years^(2)^.

Cases of accidental poisoning occur frequently in the children below 5 years of age and <1% of poisoning cases in children are serious ^(2)^. The most regular causes of accidental poisoning involve over-the-counter/prescription drugs, general domestic products, kerosene, poisonous herbs, pesticides, sting/bites by insects etc.^(1)^. In order to prevent the poisoning cases in children, parents and care takers should be educated about the potential risk factors for poisoning and methods to reduce the incidence of poisoning in children^(3)^. The duration between toxic exposure and clinical manifestations poses a chance wherein, it is crucial to reduce the uptake by neutralizing and/or removing or via using various protective agents which thwart organ injury^(2,3)^.

In Saudi Arabia, a study to study the outcomes and management of cases of poisoning in young children admitted to hospitals in Saudi Arabia^(4)^, revealed that unintentional poisoning cases in children and adults were similar in different regions., and showed that there is a requirement of public understanding on how to storage the toxic materials and the need for an urgent hospitalization in case of accidental poisoning. The study recommended strategies on careful storage of drugs in order to decrease the associated adverse events^(4)^.

Another study in Saudi Arabia, reported that young children below 12 years of age were mostly affected by intoxications (n=58, 44.2%). Drug overdose (n=119, 92.2%) was found to be the frequent reason for poisoning and the nonsteroidal anti-inflammatory and analgesic agents were mostly involved (n=26, 20.4%)^(5)^. Example of studies that reported adverse events mainly poisoning, their type of intoxication and outcomes are reported in Table 1.

**Table 1.** Example of studies that reported adverse events mainly poisoning, their type of intoxication and outcomes.

| # | Population of the study | Country of publication | Type of poison | Intent of exposure | Outcomes of poisoning | Why was this study included | References |
| --- | --- | --- | --- | --- | --- | --- | --- |
| 1 | 454,520 humans came to Poison Information & Education Centre | New Jersey, USA | Antidepressant medication | Unintentional | Adverse drug reaction resulting in hospitalization | Drug poisoning case | Vassilev, Z. P., Chu, A. F., Ruck, B., et al., (2009)a |
| 2 | 8,500 patients reported to Poison Information Center | New Jersey, USA | Cold & cough medication | Unintentional | Adverse drug reaction resulting in hospitalization | Drug poisoning case | Vassilev, Z. P., Chu, A. F., Ruck, B., et al., (2009)b |
| 3 | 1,096 cases | North Carolina, USA | Opioid analgesic | Unintentional | Deaths due to prescribed drugs | Drug poisoning causing death | Ford, M. D. (2010) |
| 4 | 2,990,000 unintended patients came to Poison Data Center | USA | Analgesic agents | Unintentional | Moderate levels of clinical effects | Drug related unintended errors | Brophy, T. J., Spiller, H. A., Casavant, M. J., et al., (2010) |
| 5 | 74 patients poisoned due to insecure heating | USA | Carbon monoxide (CO) | Unintentional | No deaths were reported | CO poisoning reports at hospital | Doğan, N. Ö., Savrun, A., Levent, S., et al., (2015) |
| 6 | 87,375 patients treated for poisoning at Poisoning Center | Egypt | Non-opioid analgesics, anti-rheumatics & antipyretics | Unintentional | 75% had minor/no effect, 119 deaths due to pesticide intake | Drug related poisoning | Azab, S.M., Hirshon, J.M., Hayes, B.D. (2016) |
| 7 | Involuntary poisoning in children reported at Center of Injury Prevention & Control | USA | Cosmetics & personal care items, pain relieving medicine, domestic cleaning items | Unintentional | 35 deaths | Poisoning caused by multiple toxicants | Schwebel, D. C., Evans, W. D., Hoeffler, S. E., et al., (2017) |
| 8 | General population including all age groups and gender | Mexico | Effluent gases, alcohol & pesticides | Unintentional | 21,700 deaths, death in most of the reported cases | Poisoning related deaths | González-Santiago, O., Morales-San Claudio, P. C., Cantu-Cardenas, L. G., et al., (2017) |
| 9 | ~750 patients at Drug & Poison Information Facility | Iran | Antimicrobials | Unintentional | Adverse drug effect (68%), hospitalization (7%) | Drug related poisoning | Sharif-Askari, F. S., Sharif-Askari, N. S., Javadi, M., & Gholami, K. (2017) |
| 10 | 207 cases within 10 months | South Africa | Organophosphate poisoning | Intentional (51%), Unintentional (22%) | Few deaths (3.4%) | Organophosphate related poisoning | Razwiedani, L. L., & Rautenbach, P. G. D. (2017) |
| 11 | 273 patients | Korean Republic | Organophosphate poisoning | Unintentional | Hospitalization | Organophosphate related poisoning | Jung, W. J., Yu, M. H., Lee, Y., et al., (2018) |
| 12 | Patients reported in Chinese National Disease Surveillance Point systems | China | Pesticides | Unintentional | 4,936 deaths | Pesticide related toxicity at National level | Wang, L., Wu, Y., Yin, P., et al., (2018) |

Most of the poisoning cases reported across the globe occur in children below the age of 5. Most of these affected children are boys^(2,4,5)^. The most prominent poisoning agent in high and middle-income countries is medicines as children accidentally consume medicines. Other poisoning agents are organic fuels, pesticides, household chemicals and carbon monoxide. In some of the Asian countries, (poisoning from bites and stings) from spiders, snakes and scorpions are also a common form of child poisoning. In Saudi Arabia, 30% of poisoning in children of 6-12 years age is caused by envenomations. Management of poison cases in various countries depend on their socio-economic conditions as high income countries have good reporting and management facilities for the patients^(6)^.

The Saudi Arabia’s Government has developed a mechanism for poison control, reporting and investigating the intoxications and drug abuse called the Poison Control and Forensic Chemistry Center, and available in eight centers across the Kingdom in every region^(7,8,9)^.

## Aims and Objectives

The aim of this study was to assess the poisoning incidences that were reported centrally via the Saudi Arabian Ministry of Health Hotline Calling Center (937), and passed to Jeddah Poison Control and Forensic Chemistry for assessment and management. The objectives of this study were:

- To identify the characteristics of poisoning in patients of different age and gender.
- To analyze the number of cases reported in various regions of the Kingdom.
- To explore the risk factors contributing to the reported poisoning incidences.
- To describe the management of cases reported to the poison control center.

## Materials & Methods

This study was conducted from 1^st^ January 2020 to 30^th^ March 2020. Data between 01-01-2019 to 30-12-2019 was collected from the Poison Control and Forensic Medical Center (PCFMC), Jeddah, Saudi Arabia. Information relevant to the “data collection form” was manually collected from archived files in the Poison Control Center. Data was collected with the help of clinical toxicologists. The clinical toxicologists in this poison center received calls from doctors in hospitals, medical centers and patients, to answer questions related to handling and treatment of poison cases. The patients included in this study were categorized on the basis of their gender and each gender was subcategorized on the basis of age. This study included cases of both acute and chronic toxicity. Factors responsible for toxicity including mode and time of poisoning were studied with respect to the variables.

The demographic variables included in this study were age, gender and region of poisoning. Various risk factors including type, pattern, place and route of poisoning were included. Various factors involved in management including any pre-hospital management, decontamination method and time of report were analyzed.

Multiple questions were included in the questionnaire to identify the identity of caller and the patient, personal history of the patient, reason for contacting the poison control center (complaint), poisoning data, general and systemic examination, routine and toxicological investigations, treatment provided, analytic report by medical toxicologist and prognosis of the condition.

Hand filled forms were collected from the PCFMC, Jeddah, Saudi Arabia. The forms were filled on the basis of calls received from different health sectors as well as public sectors via a specified phone number of the Ministry of Health by (937). This data was then typed in an Excel spreadsheet and checked for any incomplete or redundant information.

Statistical analysis of the data was done through IBM SPSS software, version 20^(10)^.

## Results

Complete information, in the form of hand-filled forms, was obtained for 253 patients from PCFMC, Jeddah. All the data obtained from forms was carefully analyzed to compile the results.

### Patients Demography

Among the total 253 patients, 198 were up to 5 years of age while the rest of patients were more than 5 years old. Mean age of the patients, including both male and female, analyzed in this study was found to be 1.2 (±0.4) years. Incidence of poisoning was found in patients of both age groups i.e. 0-5 years and above 5 years of age. However, frequency of poisoning was much higher in children up to 5 years of age than the patients above 5 years of age as shown in Table 1.

Male and female patients were (n=137, 54%) and (n=109, 43%) respectively while gender of 3% patients could not be identified. Demographic variables of all the patients included in this study were successfully analyzed as shown in Table 2

**Table 2.** Frequency and percentages of demographic variables.

| **Variables** |  | **Frequency** | **Percentage (%)** |
| --- | --- | --- | --- |
| **Age** | 0-5 years | 198 | 78 |
|  | >5 years | 55 | 22 |
| **Gender** | Male | 137 | 54 |
|  | Female | 109 | 43 |
|  | Unidentified | 7 | 3 |
| **Region of poisoning** | Eastern region | 36 | 14 |
|  | Western region | 97 | 38 |
|  | Southern region | 50 | 20 |
|  | Northern region | 18 | 7 |
|  | Middle region | 52 | 21 |

### Geographical Locations of Adverse Events Report

Cases of poisoning were reported from various regions of the Kingdom. Incidences from the eastern, western, southern, northern and middle regions were found to be 14, 38, 20, 7 and 21% of the total cases, respectively.

### Poisoning Risk Factors

This study showed that poisoning cases included both chemical and drug poisoning. However, drugs (n=155, 61% cases) were more significant risk factor than chemicals (n=92, 36%) as shown in Table 3.

**Table 3.**
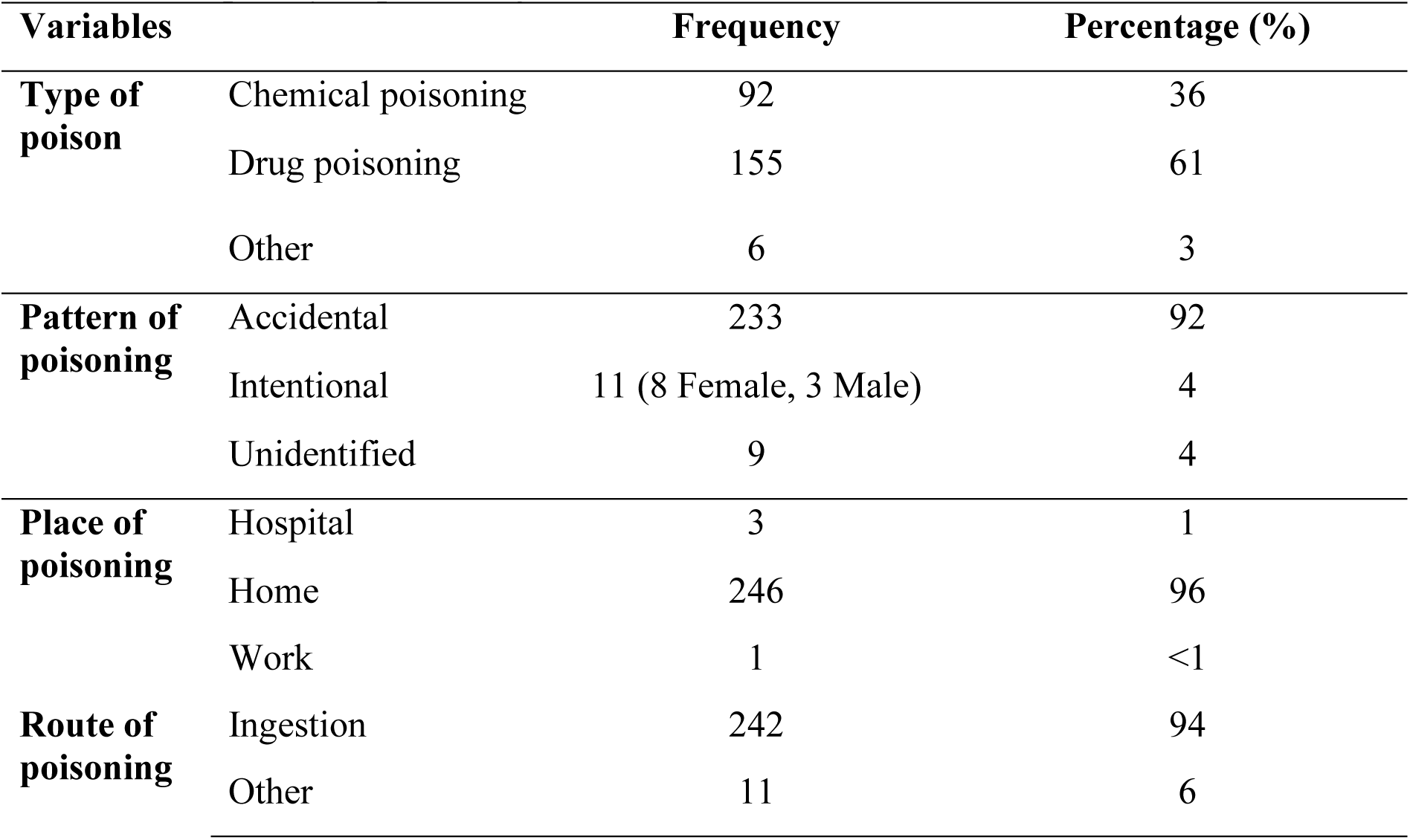
Frequency of poisoning risk factors.

For most of the cases (n=233, 92%), pattern of poisoning was accidental while only (n=11, 4%) of the cases were intentional (Table 3). Hospital or work place was not found to be the place of poisoning in most cases; most of the poisoning cases occurred at homes (96%). About 94% of the poison entered the patients through oral route or ingestion (n=242, 94%) as shown in Table 3.

Intentional poisoning was more common in females (n=8, 3%), as shown in Table 3.

### Management Consultations by the Poison Center

All of the data in this study was received from professional doctors in the poison control center. About 12% of the poisoning cases were provided with some management at homes before coming to the hospital as shown in Table 4.

**Table 4.** Management of poisoning cases at poison center.

| **Variables** |  | **Frequency** | **Percentage (%)** |
| --- | --- | --- | --- |
| **Data provider** | Doctor/Professional | 253 | 100 |
|  | Non-professional | 0 | 0 |
| **Pre-hospital management** | Absent | 220 | 87 |
|  | Present | 30 | 12 |
| **Decontamination** | Gastric lavage | 9 | 3.5 |
|  | Active charcoal | 62 | 24 |
|  | Antidote | 4 | 1.5 |
|  | No decontamination | 178 | 71 |
| **Time of reporting** | Now-1 hour | 144 | 57 |
|  | 1 hour and above | 109 | 43 |

Once the poisoning patient reached the hospital, decontamination was performed with active charcoal (n=62, 24% of the cases) or gastric lavage (n=9, 3.5% of the cases). About (n=144, 57%) of the poisoning cases were reported within an hour while 43% were reported after an hour of poisoning as shown in Table 4.

## Discussion

Poisoning affects many people across the globe thereby causing injuries or even death. This study analyzed the details of patients who suffered from poisoning and reported to the Poison Control and Forensic Chemistry Center in Jeddah, Saudi Arabia. The results of the study showed that most of the poisoning cases occurred in male children under the age of 5 years. Most of the affected children were accidentally poisoned by ingesting drugs at their homes. The poisoning cases were mostly reported within an hour to the hospital without any pre-hospital management and treated with active charcoal.

The results of the currents study showed that 78% of the cases that were reported to the poison control center were children below the age of 5 years, similar to the findings of other studies that have shown the highest prevalence of poisoning cases were among pre-school kids, particularly around the age of 2 years^(11)^. Also, previous studies showed that that children of school age group from 6 to 12 years had the lowest exposure to poison^(12)^. These results are related with those of previous epidemiologic studies on pediatric poisoning which found a bimodal age distribution of poisoning with a high incidence among toddlers^(13)^.

In the current study, poisoning cases were found to be slightly more prevalent in males (54%) than in females (43%). These results relate with previous findings which showed that in the preschool and school age groups, boys exceeded in number as compared to the number of girls^(11, 14, 15, 16, 17)^. Similarly, female cases were less common as compared to the males in the adolescent group^(15, 18, 19)^.

In this study, poisoning cases were reported from various regions of KSA. However, most of the cases were reported from Western and Southern regions while only a few cases were reported from the Northern region. This may be due to the reason that Northern region is less urbanized as compared to the Eastern and Western regions^(20)^ and parents are often not aware about how to report poison cases to the hospitals.

The current study also analyzed various poisoning risk factors, such that drug poisoning was more prevalent cause of poisoning than the chemical poisoning (Table 3). About 61% of the patients were poisoned with medicines, mainly due to accidental use of pharmaceutical agents without any prescription ^(13)^. Since most of the patients in this study are below the age of 5 years, it is evident that they can mistakenly consume drugs accidentally as shown in this study. Kids below the age of 5 are generally unable to perceive what they should eat and what not. This is the reason for excessive poisoning due to medicine in children^(21)^.

It has been found that the most common pharmaceutical compounds linked to poisoning were non-opioid analgesics in studies based in Turkey^(22)^, Palestine^(23)^, Australia^(24)^, United Arab Emirates, Hong Kong, and Saudi Arabia^(25)^. On the other hand, it has also been reported that benzodiazepines, antimicrobials and opium were the most frequently involved drugs in France^(26)^, Israel and Iran^(27)^, respectively. These variations might be brought up by the differences in the drug prescription patterns varying from country to country and country specific modifications to the packaging of the medicines and household chemicals.

Among pre-school and school age groups, the reason of the poisoning was predominantly the unintentional poisoning^(11, 14, 15, 16, 17, 28)^. However, in adults, higher rate of self-harm has been observed in adolescent females^(29)^ and it was explained by higher tendency of internalizing emotional and behavioral problems in female adolescents.

Place of poisoning was found to be home in 96% of the cases in this study. This is due to the reason that parents get unaware of their kids and the kids accidentally or unintentionally take a poison as reported previously^(30)^. Primary route of poisoning was found to be ingestion (94%) in our study as compared to other routes (6%) of poisoning like inflammation, dermal injury routes. This is in agreement with previous studies that have also shown active charcoal to be the primary method of poisoning decontamination^(31)^.

All of the data included in this study was provided by doctors or professionals that reduced the chances of any technical errors that could have been done by the non-professionals. As far as the management of poisoning cases is concerned, it was found that only 12% of the reported poisoning cases tried to do some kind of management at home while 87% were brought directly to the hospital. Most of the poisoning cases were managed in the hospital through decontamination with active charcoal (24%) as shown in Table 3. Most of the cases (57%) were reported within an hour. However, 43% of the cases were reported after one hour of poisoning. Previous studies have shown that active charcoal alone is better treatment for poisoning cases presented in hospital within one hour^(32)^. Since most of the patients were brought to hospital within 1 hour in our study, active charcoal management was the best option. This shows that the hospitals were adequately managing the poison cases.

Limitations of this study include small sample size and collection of data from only one poison control center. Greater sample size including data from extended time period, i.e., 1 year or more would have produced more detailed analysis. Despite these limitations, this study provided insight into the types and frequency of poisoning in a large city of Saudi Arabia, which merits a larger national investigation.

## Conclusion and Recommendations

The study revealed that poisoning was more prevalent in children below the age of 5, primarily males. High number of poisoning cases was reported in the Western and Southern regions while the lowest number of cases was found in the Northern region. It is concluded that most of poisoning cases were accidental and occurred at home due to ingestion of medicines. Intentional poisoning was more common in females. Most of the cases were reported within one hour of poisoning to hospital without any pre-hospital management. Prime method of decontamination at the hospitals was active charcoal.

It is recommended that parents should keep all medicines away from the reach of children, especially boys. People should be educated and special mechanisms should be developed in the Northern regions so that people can quickly report poisoning. Additionally, all the residents of the Kingdom should be educated to report the poisoning cases as soon as possible to the hospital so that chances of injury may be reduced^(33)^.

## Data Availability

Data was collected from the poisoning archived forms, between 01-01-2019 to 30-12-2019.

